# Hematological and Molecular Spectrum of Hemoglobinopathies in the Tharu Population of Nepal

**DOI:** 10.64898/2026.04.23.26351569

**Authors:** Umesh Prasad Gupta, Arati Pokharel, Kapilesh Jadhav, Indrani Jadhav, Rajendra Kumar BC, Sangam Subedi, Manish Gupta

## Abstract

Hemoglobinopathies are inherited disorders of hemoglobin, most notably sickle cell anemia and thalassemia. These conditions result from mutations in the globin genes, leading either to structural abnormalities in the globin chains or to reduced synthesis of normal globin chains. Hemoglobinopathies is a worldwide health problem according to the World Health Organization; it affects mostly the indigenous Tharu groups in Nepal. Both the global and local rates of illness and death associated with these diseases are on the rise. The objective of this study was to assess the presence of hemoglobinopathies and common mutations of the beta-globin gene within the Tharu population in western Nepal. A cross-sectional study of 1,400 Tharu individuals was conducted among individuals obtained through hospitals within the Banke district, Bardiya district, and Kailali district in western Nepal. A thorough hematological analysis was done with the use of a Sysmex XN-350 analyzer. Hemoglobin variants were detected via high-performance liquid chromatography (HPLC). The molecular characterization of the seven most common mutations of β-thalassemia was performed on a subset of 20 confirmed cases by using a real-time PCR kit.The total number of cases diagnosed with hemoglobinopathies was 14.43% (n=202 out of 1,400). Sickle cell trait (HbAS) was reported as the most prevalent type of Hemoglobinopathies (8.50% of population), followed by β-thalassemia trait (4.00%). In addition to these disorders were sickle cell disease (HbSS), HbE trait, and compound heterozygous states. Hematological parameters differed significantly across types of hemoglobinopathies, and the patterns of microcytic, hypochromic, and hemolytic anemia were also distinct. Commonly documented symptoms included fatigue and joint pain (42.5% and 23.1%, respectively). Molecular characterization of β-thalassemia cases demonstrated that most individuals were compound heterozygotes with IVS1-6 (T>C) as the most prevalent variant. The research identified that the Tharu population in western Nepal has a significant burden of hemoglobinopathies (especially sickle cell trait and β-thalassemia), highlighting the requirement for appropriate screening programs, genetic counseling and public health strategies to help manage and prevent these conditions within this particular region.

## Introduction

Hemoglobinopathies, which include sickle-cell disease (SCD) and thalassemia, are two of the most frequently encountered inherited conditions worldwide, with an estimated seven percent of the global population being carriers[1]. Once endemic to the tropical and subtropical zones, these disorders are now prevalent throughout the world due to migration[2]. Carrier screening for hemoglobinopathies can be accomplished through basic first-line testing or by referring complicated cases to a specialist. Basic first-line testing for hemoglobinopathies is done through red blood cell indices and other techniques such as High-Performance Liquid Chromatography (HPLC) or Capillary Electrophoresis (CE)[3]. To effectively manage people with hemoglobinopathies appropriately, accurate diagnoses of the particular disorder are needed for proper support and genetics counseling[4]. Sickle cell disease, one of the hemoglobinopathies, occurs with greater frequency in the western area of Nepal compared to other places within the country, and most hemoglobinopathies are found among members of indigenous Tharu community[5]. Of the several types of hemoglobinopathies, sickle cell disease (HbS) has the greatest prevalence, followed closely by thalassemia. Occasional variants are hemoglobins C (HbC), D, persistence of fetal hemoglobin (HbF), and more than 200 other types[6]. Most commonly the HbS gene is found in places once affected by Plasmodium falciparum malaria (e.g., Equatorial Africa (10 to 30%), central India (20 to 30%), eastern Saudi Arabia (up to 25%), and the Mediterranean (north Africa, Italy, Greece, and Turkey)[7]. Predictions indicate an increase in the number of hemoglobinopathies (i.e., sickle-cell disease [SCD] and thalassemia) in the future, leading to less-developed countries needing to take immediate measures to combat increased disease burden. As a result, this may lead to wider economic consequences[8]. There are also significant impacts for the Tharu ethnic group living in the western Terai region of Nepal where malaria is endemic and both SCD and thalassemia are common. Higher rates of these disorders are related to socioeconomic disadvantage and historical labour bonded backgrounds of the Tharu people[9,10]. In fact, there is insufficient research currently available regarding prevalence patterns of hemoglobinopathies in Nepal and little information is available through government agencies. Diagnostic difficulties are present in Nepal because there are no molecular testing centres located at regional hospitals and this leads to misdiagnosing (alpha-thalassemia misdiagnosed as iron deficiency anaemia), resulting in ineffective treatment/recovery. While there has been recent establishment by the government to offer financial support to persons diagnosed with SCD, there is no registry of those affected or carriers; therefore the availability and delivery of such government funded services will be limited[11]. While hemoglobinopathies mainly afflict the Tharu group, they do occur outside of this ethnicity and through other high-population areas, namely Pokhara and Kathmandu[12]. Enhanced diagnostic capabilities and greater understanding of these hereditary diseases can drastically improve the overall health burden on Nepalis suffering from these blood disorders[9,10]. The objectives of this study are: to ascertain the prevalence (frequency) and burden (impact) of *β* -Thalassemia; to characterise the spectrum of mutations and relative frequency (type of mutation), to analyze and compare the hematological parameters of the population to determine the severity of *β* - Thalassemia in those affected; to evaluate the demographic distribution of *β* - Thalassemia between males and females with respect to age; to provide a detailed record of all patients with hemoglobinopathies and their clinical presentation; and to define the genetic basis and mutation spectrum associated with *β* -Thalassemia.

## Materials and Methods

### Study Design and Setting

The study was performed over a four-year period (2018-2022). Sample collection was started from May 2019 and ended on November, 2022 at three locations in western Nepal: the three sample collection sites used to obtain specimens were Nepalgunj (included Dang, Banke, Bardiya) and Dhangadhi (Kailali). There were two specific locations designated for the collection of samples—Nepalgunj and Dhangadhi—wherein samples from three of the four districts were collected at Nepalgunj (Dang, Banke, Bardiya) and those from Kailali were obtained at Dhangadhi.

### Study Population

A total of 1,400 Tharu people who received treatment at the Bheri Hospital in Nepalgunj and the Seti Provincial Hospital in Dhangadhi participated, of whom 432 were recruited from Banke District, 599 from Bardiya District, and 369 from Kailali District. Participants were both genders and represented a broad age range. Participants above the age of 2, who had signs of or were suspected to have anemia, malaise, fever, or clinically suspect Hemoglobinopathies, according to their personal or family medical history, were eligible for inclusion. Non-Tharu participants were excluded and those with a recent history of receiving a blood transfusion were tested for eligibility.

### Ethical Considerations

Written Informed consent was obtained from all participants through signature or thumb impression. Demographic data, clinical information and family history of hemoglobinopathies were collected via a structured survey questionnaire. Ethical approval was obtained from the Institutional Review Committee (IRC) and Pokhara University Research Center (PURC), Pokhara University, Nepal (Ref No 171/075/76), dated May 16, 2019.

### Sample Collection and Transportation

Three milliliters (3 mL) of venous blood were collected in EDTA (ethylene diamine tetra acetic acid) vacutainer tubes from each subject using aseptic methods. Samples were kept at 4C in a cold chain box and then transported to the Central Diagnostics Laboratory (CDL) located in Kathmandu, Nepal, which is classified as a Grade A reference laboratory, where they will undergo hematological and molecular analysis.

### Laboratory Methods

Hematology tests included the following performed by the Sysmex XN-350 Hematology Analyzer: RBC count; hemoglobin concentration; hematocrit (HCT); reticulocyte count; and red cell indices (mean corpuscular volume (MCV), mean corpuscular hemoglobin (MCH), and mean corpuscular hemoglobin concentration (MCHC)). The internal quality control sample of cell pack was analyzed prior to the start of each batch of test samples.

The Bio-Rad D-10 HPLC system (Bio-Rad, USA) was used to identify hemoglobin variants. The concentrations of HbA2, HbF and non-functional hemoglobins were determined. Daily calibration and internal quality controls were performed according to manufacturer’s instructions.

Twenty Hemoglobinopathies positive samples that were HPLC confirmed were selected randomly for β-globin gene mutation testing. The HELINI Human Blood DNA Mini Spin Prep Kit (HELINI Biomolecules, India) was used to extract genomic DNA from these samples. All samples were assessed for DNA quality and concentration spectrophotometrically and were subsequently frozen at -20 °C prior to testing.

The HELINI β-Thalassemia Real-time PCR Kit (HELINI Biomolecules, India) was used for Mutation detection. The seven (7) most common beta-thalassemia mutations (IVS1-1(G/A); IVS2-1(G/A); IVS1-110(G/A); IVS1-5;IVS1-6 (T/C); Codon 8/9(+G); Codon 41/42(-TCCT) using PCR with Denaturation @ 95°C / Annealing @ 60°C / Extension @ 72°C X 35 cycles. Fluorescent determination was performed by means of FAM dye and analyzed according to the kit manufacturer.

## Results

### Study Population Overview

Amongst 1,400 Tharu individuals in western Nepal, two hundred and two (14.43%) were found to carry a Hemoglobinopathies using HPLC although most (1,198; 85.57%) were found to be non-carriers of a Hemoglobinopathies (Fig 1). The following other hemoglobinopathies were also found in the study sample: sickle cell homozygous (HbSS, 4.45%); HbE heterozygous (3.96%); compound heterozygous for HbS and β-thalassemia (1.98%); hereditary persistence of fetal hemoglobin (HPFH) trait (1.98%); delta-beta thalassemia trait (0.99%). Based on these results, one can see that there is a wide variety of Hemoglobinopathies in the Tharu population in western Nepal.

**Fig 1.**
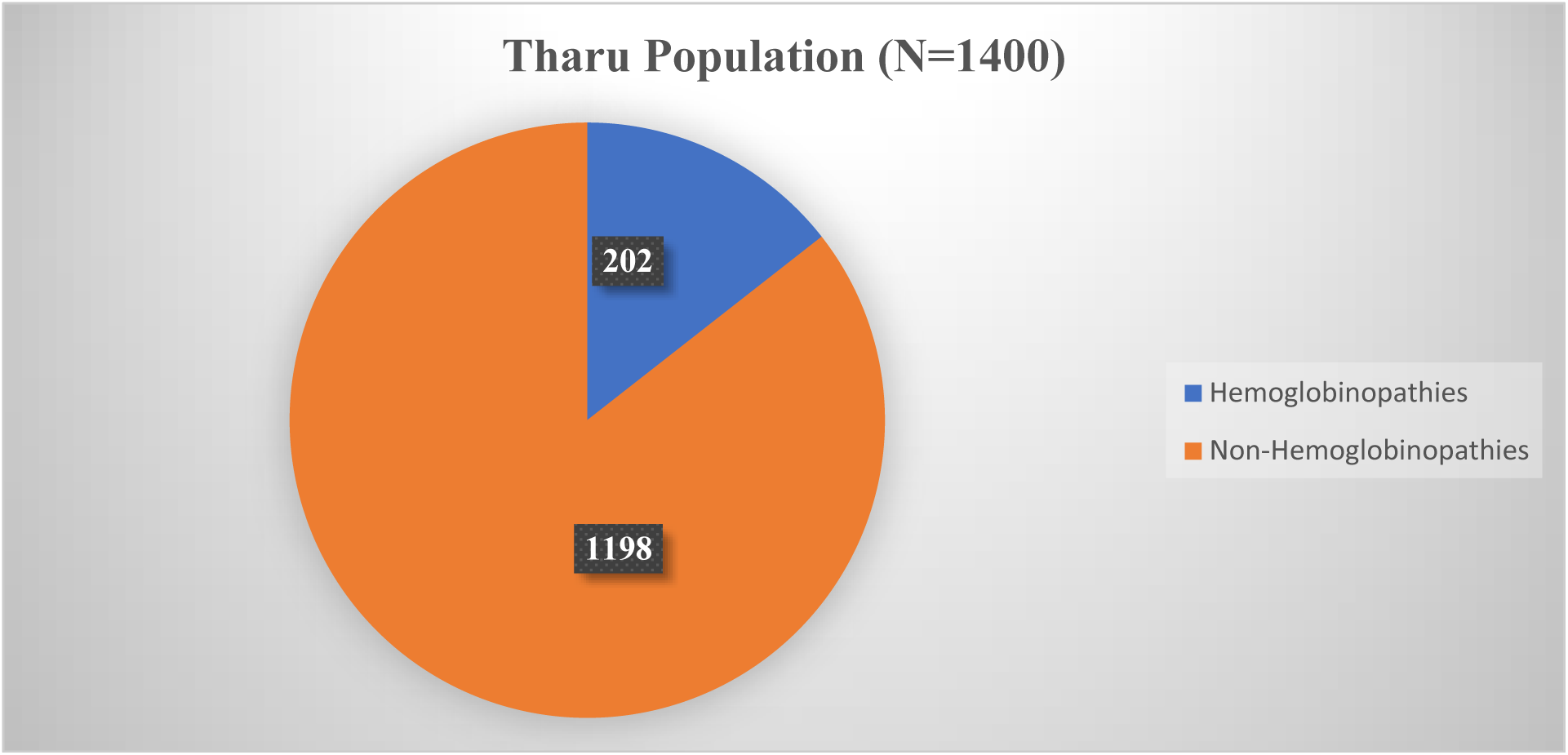
Prevalence of Hemoglobinopathies.

### Spectrum and Prevalence of Hemoglobinopathies

The analysis of the complete sample population using HPLC shows that the greatest number of individuals with a Hemoglobinopathies from the sample population had sickle cell trait (HbAS), which accounted for fifty-eight point ninety-one percent of individuals with a Hemoglobinopathies, while twenty-seven point seventy-two percent of the individuals had a β-thalassemia trait (Table 1). The following other hemoglobinopathies were also found in the study sample: sickle cell homozygous (HbSS, 4.45%); HbE heterozygous (3.96%); compound heterozygous for HbS and β-thalassemia (1.98%); hereditary persistence of fetal hemoglobin (HPFH) trait (1.98%); delta-beta thalassemia trait (0.99%). Based on these results, one can see that there is a wide variety of Hemoglobinopathies in the Tharu population in western Nepal.

**Table 1.** Distribution of Hemoglobinopathies among the Tharu Population (n = 1,400)

| Hemoglobinopathies | Number of Cases | Percentage of Hemoglobinopathies (%) | Percentage of Total Population (%) |
| --- | --- | --- | --- |
| Sickle cell trait (HbAS) | 119 | 58.91 | 8.50 |
| β-Thalassemia trait | 56 | 27.72 | 4.00 |
| Sickle cell homozygous (HbSS) | 9 | 4.45 | 0.64 |
| HbE heterozygous | 8 | 3.96 | 0.57 |
| Compound heterozygous<br>(HbS/β-Thalassemia) | 4 | 1.98 | 0.29 |
| HPFH trait | 4 | 1.98 | 0.29 |
| Delta-beta thalassemia<br>trait | 2 | 0.99 | 0.14 |
| Total hemoglobinopathies | 202 | 100 | 14.43 |
| Non-Hemoglobinopathies | 1,198 | – | 85.57 |

### Hematological Profile Across Disorders

The evaluation of Complete Blood Cell Counts (CBC) among 14000 samples reveals that there are highly significant differences in the hematological parameters of non-hemoglobinopathies vs. hemoglobinopathies. The largest number of RBCs was found in the population of non-hemoglobinopathies followed by delta β-thalassemia trait. Moderate decreases in RBC counts were observed for HbE heterozygous while both hemoglobin and hematocrit were lower than normal for β-thalassemia trait and HPFH trait. WB, MCV, MCH and MCHC and RDW are consistently higher in non-hemoglobinopathies than in hematologic disorders. Moderate decreases were also found in Hb and HCT of both sickle cell trait and HbE heterozygous than those found in β-thalassemia trait, HPFH trait, sickle cell homozygous, compound heterozygous for HbS and β-thalassemia and delta β-thalassemia trait which had significantly lower levels of Hb and HCT than normal. In comparison, individuals with HPFH trait had the greatest MCH, MCHC, and least RDW; sickle cell homozygotes presented with increased MCV, MCH, and MCHC values. Among all groups, those who were compound heterozygous for HbS and β-thalassemia had the highest RDW. Table 2 presents the hematological parameters for various types of hemoglobin disorders showing systemic median (interquartile range) values, with significant differences in their RBC levels (p < 0.05) when compared to non-hemoglobinopathies, sickle cell trait (HbAS), and β-thalassemia. Sickle cell homozygous (HbSS) individuals had a significantly reduced RBC median compared to non-hemoglobinopathies but a higher MCV and MCH. Hematology testing (RBC, hemoglobin, hematocrit, MCV, MCH, MCHC, and RDW) was performed for each of these groups of people with different types of hemoglobinopathies (i.e., sickle cell trait, sickle cell homozygous, β-thalassemia trait). There was variance in hematological parameters for each group, with sickle cell homozygous individuals having reduced levels, in particular hemoglobin and hematocrit level, compared to other groups. In addition, sickle cell homozygous and β-thalassemia trait individuals had significantly lower levels of hemoglobin than individuals with the normal hemoglobin trait (i.e., individuals without a Hemoglobinopathies). Of the three hemoglobinopathies studied, sickle cell homozygous had the highest levels of MCV and MCH.

**Table 2.** Hematological Parameters Across Different Hemoglobinopathies.

| Hemoglobin Disorder | RBC |  | HGB (g/dL) |  | HCT (%) |  | MCV (fL) |  | MCH (pg) |  | MCHC (g/dL) |  | RDW |  |
| --- | --- | --- | --- | --- | --- | --- | --- | --- | --- | --- | --- | --- | --- | --- |
|  | (×10<sup>12</sup>/L) |  |  |  |  |  |  |  |  |  |  |  |  |  |
|  | Median |  | Median |  | Median |  | Median |  | Median |  | Median |  | Median |  |
|  | (IQR) |  | (IQR) |  | (IQR) |  | (IQR) |  | (IQR) |  | (IQR) |  | (IQR) |  |
| Non-hemoglobinopathies | 5.01 | (4.70–5.35) | 11.10 | (10.40–12.00) | 37.30 | (35.30–39.70) | 74.55 | (70.00–80.32) | 22.10 | (20.45–24.40) | 29.70 | (29.10–30.40) | 13.20 | (12.60–14.00) |
| Sickle Cell Trait (HbAS) | 4.85 | (3.98–5.22) | 10.80 | (9.40–11.90) | 34.00 | (28.45–37.35) | 71.60 | (68.70–76.20) | 22.90 | (20.45–25.90) | 30.20 | (29.30–31.25) | 14.10 | (13.10–19.20) |
| β-Thalassemia Trait | 4.48 | (4.12–4.88) | 9.85 | (9.00–10.77) | 31.00 | (28.93–33.73) | 70.25 | (69.00–74.55) | 21.00 | (19.80–22.00) | 29.20 | (28.50–29.70) | 13.70 | (13.00–14.30) |
| Sickle Cell Homozygous (HbSS) | 4.21 | (3.95–5.18) | 10.75 | (7.72–13.32) | 31.10 | (23.93–41.70) | 78.25 | (75.67–81.82) | 25.95 | (23.55–28.40) | 30.45 | (28.75–31.18) | 14.25 | (12.77–15.97) |
| HbE Heterozygous | 4.87 | (3.73–5.33) | 9.85 | (9.20–11.07) | 31.10 | (29.28–33.23) | 69.74 | (66.02–80.35) | 21.30 | (20.03–24.28) | 28.80 | (29.45–30.68) | 14.15 | (13.55–14.40) |
| Compound HbS–β-Thalassemia | 3.93 | (3.75–4.36) | 9.20 | (8.37–11.30) | 33.30 | (29.05–38.15) | 78.55 | (69.55–86.80) | 22.50 | (20.60–27.70) | 27.80 | (25.53–28.88) | 18.00 | (16.47–20.50) |
| HPFH Trait | 4.05 | (3.90–4.72) | 10.40 | (9.35–11.60) | 32.00 | (28.25–37.78) | 73.00 | (69.25–78.40) | 21.00 | (17.50–23.90) | 28.00 | (26.25–30.13) | 12.55 | (12.05–15.15) |
| Delta β-Thalassemia Trait | 5.42 |  | 9.95 |  | 33.65 |  | 62.10 |  | 18.30 |  | 29.50 |  | 14.05 |  |

### Gender-wise Distribution of Hemoglobinopathies

HPLC analysis of 202 cases of confirmed Hemoglobinopathies showed that there were a higher number of females (132 cases) than males (70). Fig 2 shows that sickle cell trait was the most common Hemoglobinopathies in both groups, occurring in 76 females and 43 males. β-thalassemia trait was also more common in females (38 cases) than males (18 cases). Sickle cell homozygous, HbS and β-thalassemia compound heterozygous, HbE heterozygous and β-thalassemia variants were detected less often than the previous variants. It was interesting to note that only female patients had delta beta thalassemia trait or hereditary persistence of fetal hemoglobin (HPFH) trait.

**Fig 2.**
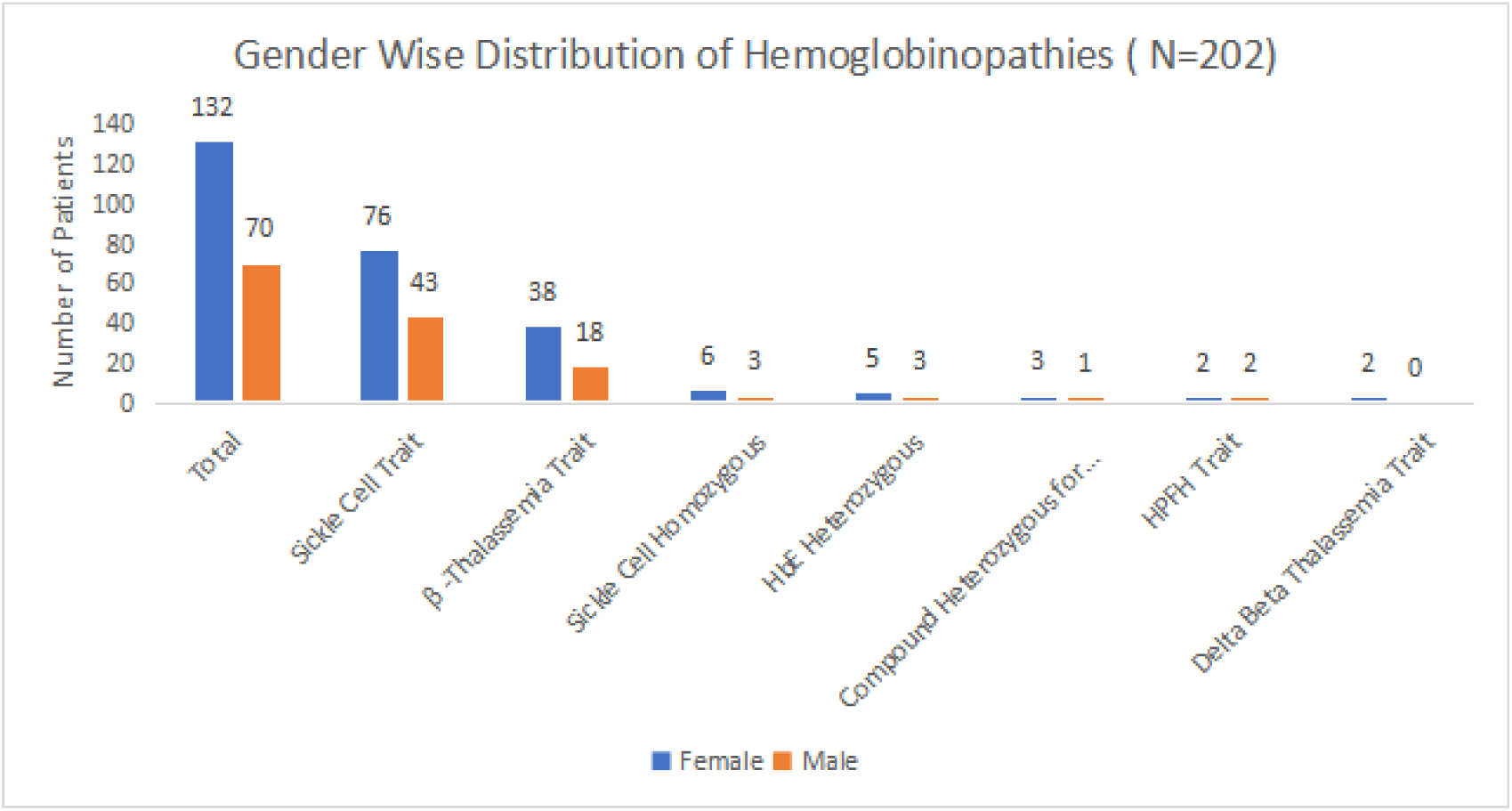
Gender-wise distribution of hemoglobinopathies subjects.

### Age-wise Distribution of Hemoglobinopathies

Analysis of 202 cases of Hemoglobinopathies revealed that the sickle cell trait is by far the most common, especially in the age groups of 13-30 years (54) and 2-12 years (48). The second most prevalent was β-thalassemia trait with a greater number of cases also found in the 13-30 (29) and 2-12 (22) years, age in brackets. Less common disorders, such as sickle cell disease (homozygous) and HbE (heterozygous), were mostly reported in the 13-30 age group (Fig 3). Rare variants, including the compound heterozygous state for HbS and β-thalassemia and the HPFH trait, were found only in youth, while the delta β-thalassemia trait, which was sporadic among all individuals, occurred most often in children. Overall, the incidence of hemoglobinopathies dramatically decreases in adults over 50 years of age.

**Fig 3.**
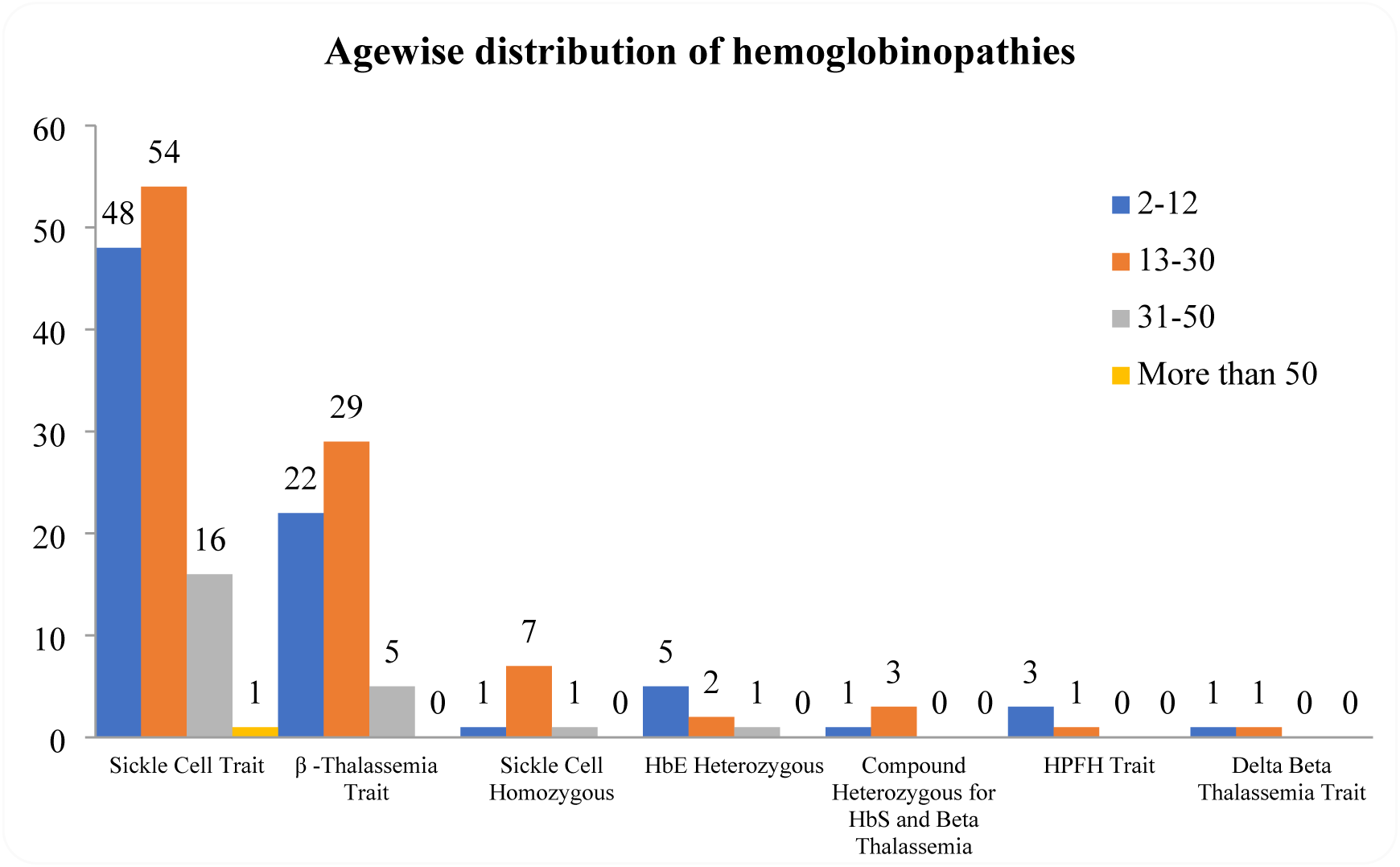
Age-wise distribution of hemoglobinopathies subjects.

### Clinical Symptomology

Fatigue was the most recorded complaint in 202 patients with a Hemoglobinopathies and 42.5% (n = 86) reported having fatigue. The next most recorded symptom was joint pain (23.1% of total patients = 47) but those were much less frequent than fatigue. Approximately, 12.1% (n = 24) and 11.2% (n = 23) requested a blood test for pallor and shortness of breath respectively. The following symptoms were also reported much less frequently: fever (5.4%, n = 11), jaundice (3.4% n = 7) and eye symptoms (n = 2.3%). Therefore, both fatigue and joint pain were predominant symptoms of those studied while the remaining symptoms were reported to occur at substantially lesser rates (Fig 4).

**Fig 4.**
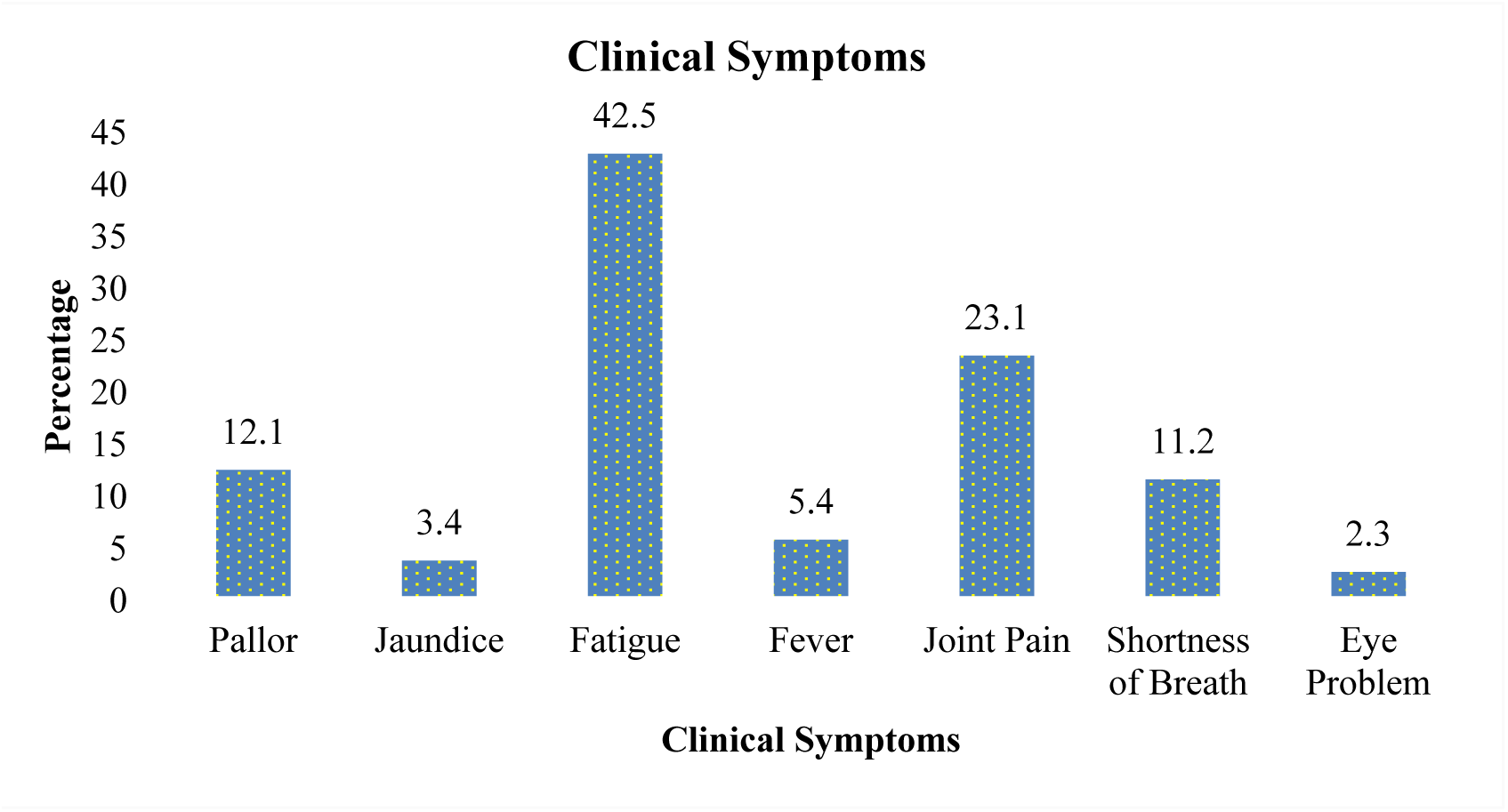
Clinical symptoms among hemoglobinopathies subjects.

### Molecular Basis of *β*- Thalassemia

The variation in mutations within the identified confirmed β-thalassemia Patient Population from the Confirmed Tharu Population (n = 20) has been demonstrated by Fig 5. Among the samples confirmed to be β-thalassemic, the majority of individuals were compound heterozygotes for gene mutations. The most frequent variant identified within this study was IVS-6, which was detected in the heterozygous state only. The amounts of compound heterozygosity seen for the IVS-1, IVS-2, and IVS-5 mutations were 15/20, 18/20, and 19/20 patients that showed compound heterozygosity, 4/20 patients had homozygous mutations for the IVS-1 mutation and 1/20 had a homozygous mutation for the IVS-2 mutation. The other mutations (IVS-110, Codon 8/9 and Codon 41/42) were found only as compound heterozygous in patients while there were 1/20, 9/20 and 17/20 cases that did not have a mutation for these same loci.

**Fig 5.**
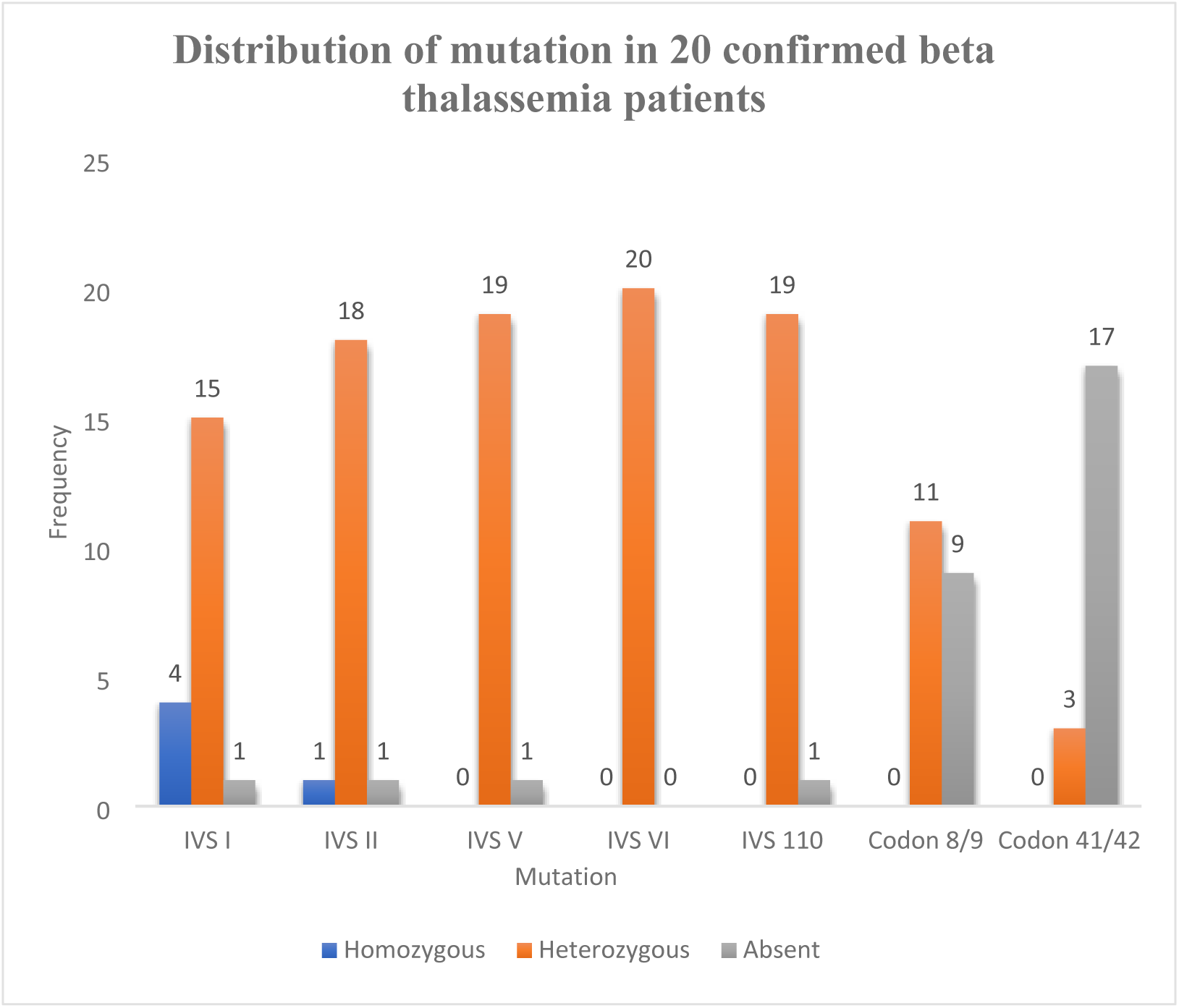
Distribution of Heterozygous and Homozygous mutation in 20 confirmed beta thalassemia patients.

Overall, the vast majority of β-thalassemia cases (close to 100%) were heterozygous mutations, while there were only five samples with specific homozygous mutations. The most common heterozygous mutations were IVS-6 and IVS-110 (19% each), followed by IVS-5 (18%), IVS-2 (17%), IVS-1 (14%), codon 8/9 (10%) and codon 41/42 (3%).

## Discussion

The Tharu people who live in the western Terai districts of Banke, Bardiyā, and Kailali are located in an area that has a long history of being a malaria endemic region and possessing a high prevalence of hemoglobinopathies[5,13].1,400 blood samples were taken from those regions, and the total prevalence of hemoglobinopathies in the Tharu people was 14.43%, indicating that the presence of hemoglobinopathies in the Tharu population is a major public health issue. Of all recognized hemoglobin disorders, the most frequently found disorder was sickle cell trait, which made up 58.91% of all hemoglobin disorders and 7.58% of People’s incidence in trials. Conversely, the homozygous sickle cell disease represented only 4.45% of hemoglobin disorders. This distribution is consistent with the global incidence pattern, with sickle cell trait being found at much higher rates than sickle cell homozygous disease, due to the advantage given to carriers who were immune to Malaria and had lower mortality rates than individuals with severe disease[1,14]. There are reports of similar rates of sickle cell trait in the Terai regions of Nepal and bordering areas of India between 14-30%, among some ethnic and tribally based populations[15,16]. Sickle cell trait is typically asymptomatic but carries a significant public health burden due to its prevalence in the population. Parents with sickle cell trait have a 25% chance of having children with sickle cell disease (HbSS), emphasizing the need for population screening and genetic counseling for those at greatest risk of having the disease.

β-Thalassemia trait was the second most frequently diagnosed Hemoglobinopathies, with a frequency of β-Thalassemia trait among our cases of 27.72%. Those individuals with β-thalassemia trait typically either present with mild anemia or have no clinical symptoms at all; however, the high frequency of carriers creates a high likelihood of transmission of the disease to subsequent generations. The continued occurrence of β-thalassemia trait in areas where malaria is endemic can likely be attributed to its protective effect from developing malaria, similar to SCT[1]. The prevalence found in this study agrees with previous studies and data from the Terai region of Nepal and other previous studies in India[5,17].

Approximately 3.96% of patients with hemoglobinopathies have HbE heterozygous traits. HbE is mostly found in Southeast Asian countries (e.g., Thailand, Cambodia, and Bangladesh), but the finding of HbE in the western part of Nepal is an indicator that the origins of east and west parts of South and Southeast Asia share some common history through genetic fluidity from migration over time18. HbE heterozygotes have mild anemia and fewer clinical symptoms. However, the presence of HbE along with beta-thalassemia may lead to meaningful clinical implications. The same prevalence of HbE may be found in people living close to the Indian border and in eastern Nepal[18].

1.98% of people diagnosed with Hemoglobinopathies display Compound Heterozygosity for HbS and Beta-thalassemia1. The clinical severity of this condition varies significantly, depending on the specific Beta-thalassemia mutation involved[19]. Some patients may present with symptoms consistent with Sickle Cell Disease but have relatively higher amounts of fetal hemoglobin (HbF) than patients with SCD, which could potentially reduce symptoms; nonetheless, patients continue to experience considerable clinical severity due to conditions caused by iron deficiency and/or disease. Additionally, the presence of patients with compound heterozygous hemoglobins further illustrates the complexity of Hemoglobinopathies in these patients, highlighting the importance of accurate molecular diagnosis and informative genetic counseling[20].

Two rare Hemoglobinopathies types, the Hereditary Persistence of Fetal Hemoglobin (HPFH) trait (1.98%) and delta β-thalassemia trait (0.99%), were also found. Both of these conditions are associated with increased HbF levels. HPFH is usually benign, and does not produce clinically significant anemia, while a patient with delta β-thalassemia may demonstrate milder anemia that could be misdiagnosed as having a β-thalassemia trait due to similarities in their hematological features[21]. The presence of these two variants confirms the genetic diversity within the Tharu people.

The primary diagnostic instrument in this study was high-performance liquid chromatography (HPLC) because it provides reliable and fast results, and it can distinguish different types of hemoglobins from one another. Different types of hemoglobin showed up differently when comparing samples between hemoglobinopathies. In the β-thalassemia trait, for example, the HbA₂ level is elevated, while in the HPFH, delta β-thalassemia and compound heterozygotes, the HbF level is increased. Conversely, the predominant hemoglobin in sickle cell disease and the compound heterozygotes is hemoglobin S. Therefore, these HPLC findings corroborate previously reported studies from Africa, India, and elsewhere to support the diagnostic use of HPLC for population-based screening programs[17,22].

Molecular analysis indicates major regional differences in β-thalassemia mutations. Most frequently identified mutations were IVS6 (T>C) and IVS1-110 (G>A), which together accounted for 38% of all mutations. IVS6 (T>C) was restricted to western Nepal and has not previously been described as one of the major mutations in patients from central or eastern Nepal, India, or any global databases[1,23]. However, IVS1-110 (G>A) has been reported consistently across southern Asia including Nepal, suggesting VII highly distributed throughout southern Asia. Other mutations included: IVS1-5 (G>C), IVS2 (G>A), codon 8/9 (+G), and codon 41/42 (−TCCT) at lower frequencies, which reinforces the need for regionally specific mutation panels for accurate molecular diagnosis[23].

The gender analysis indicates that females made up the higher proportion of participants (the male:female ratio was 1.89:1. It is likely that this imbalance results from men migrating for work during the study and has been documented in other studies conducted in Nepal[24]. Other potential reasons include: differences in how men and women seek health care and differences between men and women accessing cancer screenings.

From the age distribution of hemoglobinopathies it is evident that the most prevalent age group included individuals aged 13-30 years with the second largest number of Hemoglobinopathies cases being reported to occur in individuals aged between 2-12 years of age. The overwhelming majority of Hemoglobinopathies cases (both sexes combined) are between the ages of 2-30, this is consistent with the findings of similar studies in the Terai region of Nepal and Southeast Asia[23,25]. The fact that Hemoglobinopathies cases predominately occur between the reproductive ages illustrates the need for early diagnosis and the provision of genetic counselling. With respect to geographical distribution of Hemoglobinopathies cases, Bardiya district had the highest percentage of Hemoglobinopathies (40%) compared to Kailali (32.67%) and Banke (27.33%). This geographic distribution is consistent with other factors such as population density, history of malaria endemics, and findings from prior studies in the same region.

The majority of patients reported having fatigue as a symptom of their condition, whereas symptoms such as joint pain, pale skin, or feeling out of breath came next. These same symptoms are often seen in those with chronic anemia due to the breakdown of red blood cells associated with sickle cell disease/Hemoglobinopathies[26,27]. Blood counts have shown various findings based on the type of Hemoglobinopathies; therefore, assessing the degree of hemoglobin and hematocrit helps in determining the presence of serious illnesses. Sickle cell homozygous and compound heterozygous (HbS and β-thalassemia) conditions were found to have very low levels of hemoglobin and hematocrit, while patients with a β-thalassemia trait had normal red blood cell size but decreased mean corpuscular volume (MCV) and mean corpuscular hemoglobin (MCH). Compound heterozygous individuals had a very high RDW value, indicating a considerable degree of variability when measuring the size of red blood cells (i.e., the red blood cells being significantly larger or smaller compared to each other)[28,29].

## Conclusion

Among the Tharu individuals, 14.43% have been found to be affected by hemoglobinopathy diseases caused largely by sickle cell disease and β-thalassemia traits (8.50% and 4.00% respectively). The various disorders have different hematologic characteristics resulting in different patterns of anemia, with tiredness and arthralgia being the two most recognised clinical symptoms amongst this group. In molecular studies of β-thalassemic individuals, the major mutation was IVS1-6 (T>C). The majority of the individuals are found to have compound heterozygous mutations responsible for the hemoglobinopathies. This illustrates the marked degree of genetic diversity for targeted screening programs and genetic counselling services in the Tharu population. This emphasizes the need for screening programs and genetic counselling services.

## Data Availability

All relevant data underlying the findings of this study are fully available within the manuscript and its Supporting Information files. Additional anonymized datasets generated and analyzed during the current study are available from the corresponding author upon reasonable request, in compliance with institutional ethical guidelines.

## Acknowledgments

We sincerely acknowledge the invaluable support and collaboration provided by the School of Health and Allied Sciences, Pokhara University; the Central Diagnostic Laboratory, Kathmandu; Bheri Zonal Hospital, Nepalgunj; and Seti Provincial Hospital, Dhangadhi. Their contributions were instrumental in facilitating this research. We further extend our gratitude to the University Grants Commission for its partial financial assistance.

